# Effect of Adding Bowel Ultrasound to Standard Radiography for Suspected Necrotizing Enterocolitis: A Randomized Clinical Trial

**DOI:** 10.64898/2026.01.07.26343610

**Authors:** Alain Cuna, Sarah Foster, Maura Sien, Allison Scott, Laura Gillis, Jill Jones, Sherwin S. Chan

## Abstract

**Importance:** Bowel ultrasound (BUS) offers real-time assessment of bowel integrity and perfusion, but its clinical impact on diagnosis and management of necrotizing enterocolitis (NEC) remains unclear.

**Objective:** To evaluate whether adding BUS to standard abdominal radiography (AXR) improves diagnostic decision-making and clinical outcomes in infants evaluated for suspected NEC.

**Design:** Randomized clinical trial.

**Setting:** Level IV neonatal intensive care unit (NICU) of a freestanding children’s hospital (BUS-experienced) and a level III NICU of an adult hospital (BUS-naïve).

**Participants:** Infants undergoing imaging for suspected NEC.

**Interventions:** Infants randomized by calendar month to AXR alone or AXR + BUS. Clinical management followed standard NICU practice.

**Main Outcomes and Measures:** The primary outcome was time to full enteral feeds within 30 days of NEC concern. Secondary outcomes included duration of antibiotics and bowel rest. Analyses followed the intention-to-treat principle, with per-protocol sensitivity analyses. Post-imaging survey assessed the perceived impact of adding BUS on diagnostic and therapeutic decision-making.

**Results:** A total of 169 infants (199 NEC evaluations; 114 AXR, 85 AXR + BUS) were included; 84% were preterm, and 62% were from the level IV NICU. Among infants ultimately classified as *extended NEC rule-out* (NEC excluded after repeated imaging evaluations), BUS was associated with faster recovery to full feeds (median 4 vs 8 days; hazard ratio [HR], 1.89; 95% CI, 1.23–2.91). No differences were observed among infants with *quick NEC rule-out* (NEC excluded after single imaging evaluation) or *confirmed NEC*. BUS did not prolong antibiotic or bowel rest duration. Per-protocol analysis yielded similar results for all outcomes. Neonatologists reported greater diagnostic and therapeutic confidence with BUS.

**Conclusions and Relevance:** In this dual-site randomized trial, adding BUS to standard AXR accelerated feeding recovery among infants with prolonged diagnostic evaluations for suspected NEC, without increasing adverse events. BUS may enhance diagnostic confidence and streamline management in cases of diagnostic uncertainty. Larger multicenter studies are warranted to confirm these findings and guide implementation across NICU settings.

**Trial Registration:** ClinicalTrials.gov Identifier NCT05573113

**Key Points:** *Question:* Does adding bowel ultrasound to standard radiography improve outcomes among infants evaluated for suspected necrotizing enterocolitis?

*Findings:* In this randomized diagnostic trial of 169 infants, adding bowel ultrasound shortened time to full feeds among infants with prolonged diagnostic evaluations without increasing adverse events.

*Meaning:* Bowel ultrasound may enhance diagnostic confidence and expedite feeding recovery in infants with diagnostic uncertainty for NEC.

## INTRODUCTION

Necrotizing enterocolitis (NEC) is a devastating inflammatory bowel disease primarily affecting preterm infants but also term infants, with a high risk of morbidity and mortality.^1–6^ Imaging plays an important role for the diagnosis of NEC, yet current imaging modalities, particularly abdominal radiography (AXR), have notable limitations in sensitivity, specificity, and variability in interpretation.^7–9^ Bowel ultrasound (BUS) has emerged as a promising adjunct imaging tool, offering real-time assessment of bowel perfusion, wall integrity, and pneumatosis.^10–14^ Meta-analyses suggest that BUS enhances NEC diagnosis and may improve risk stratification and prognostication.^15–17^

Despite growing interest in BUS for NEC, its impact on patient outcomes remains unclear. Current studies are largely observational, often single-center, and lack standardized protocols, leading to variability in interpretation and implementation.^18–23^ In Fryback and Thornbury’s hierarchy of diagnostic efficacy, such studies address only diagnostic accuracy (Level 2) – how well an imaging test can detect disease.^24^ Importantly, no prospective randomized trials have evaluated whether BUS improves NEC diagnosis (Level 3), management (Level 4), and patient outcomes (Level 5) compared with standard AXR alone.^25^ Another critical gap is that most studies have been conducted in children’s hospitals with pediatric radiologists, raising concerns about the generalizability of BUS in level III neonatal intensive care units (NICUs) within adult hospitals, where radiology expertise and resource availability may differ. Addressing these gaps is essential to determine whether BUS should be integrated into routine neonatal care across diverse clinical settings.^26–28^

To address these gaps, we conducted a randomized diagnostic trial evaluating the impact of adding BUS to standard AXR for NEC evaluation in two different settings: a level IV NICU in a freestanding children’s hospital, and a level III NICU in an adult hospital. The primary hypothesis of our study was that adding BUS would enhance NEC diagnostic accuracy and clinical decision making and this would be manifest by decreased number of days to full feeds for infants with suspected NEC compared to AXR alone. Our secondary analyses measured whether BUS enhances clinical diagnostic decision-making, influences management strategies.

## METHODS

### Study Design and Setting

This was a comparative effectiveness randomized diagnostic trial evaluating the effect of adding BUS to standard AXR for infants with suspected NEC (Clinicaltrials.gov NCT05573113). The Children’s Mercy IRB approved the study (STUDY00002293) and granted a waiver of consent based on both clinical paradigms being standard of care, no more than minimal risk, no adverse impact on subject rights, and impracticability of conducting the study without the waiver. The trial was conducted at an 87-bed level IV NICU (Children’s Mercy Hospital) and a 34-bed level III NICU (University of Kansas Medical Center). BUS was available at the level IV NICU since 2018 but used variably. BUS was not available at the level III NICU until participation in the study. Both sites used a standardized BUS protocol and reporting template (**eTable 1 and eTable2 in the Supplement**).^25,29^

### Study Subjects and Intervention

Trial included infants with suspected NEC requiring imaging for further evaluation. Infants with gastrointestinal anomalies that limit BUS (ie, gastroschisis, omphalocele) were excluded. Randomization was by calendar month when infant was born: odd months to AXR only, even months to AXR plus BUS. Infants in the AXR group underwent standard portable supine AXRs, with additional left lateral decubitus or cross table views as needed. Infants in the AXR plus BUS group underwent standard AXRs with study BUS consisting of grayscale, color Doppler, and spectral Doppler images supplemented with cine acquisitions in transverse and sagittal planes.^29^ Cross-over between groups was discouraged but allowed per neonatologist’s discretion.

### Trial Treatments and Procedures

Any repeat imaging for continued NEC concern or follow-up was also left to the neonatologist’s discretion and consisted of the imaging modality/modalities to which infants were randomized. All AXR and BUS examinations were performed and available for review as per usual clinical practice. The diagnosis of NEC was made by the treating neonatologist based on interpretation of infant’s clinical features, laboratory data, and imaging results. The treatment of NEC, including duration of antibiotics and bowel rest, was also left to the discretion of the clinical team. In general, medical NEC was treated with antibiotics and bowel rest for 7-10 days, while surgical NEC was treated for 14 days, in addition to laparotomy or peritoneal drainage.^30,31^ Because the modified Bell staging criteria^30,31^ do not include BUS findings, we used a pragmatic, treatment-based classification to categorize infants into 3 diagnostic cohorts based on their clinical course: *NEC Ruled In*, *Quick Rule-Out*, and *Extended Rule-Out*. *NEC Ruled In* included infants treated with ≥7 days of antibiotics and bowel rest for NEC, while *Quick Rule-Out and Extended Rule-Out* referred to infants for whom NEC was excluded after single or repeated imaging, respectively. Classification was determined by chart review, with clarification from the treating neonatologist when cases were ambiguous. Neonatologists completed a survey ∼24 hours after NEC concern to assess the perceived impact of imaging on clinical decision-making (**eTable 3 in the Supplement**).

### Outcomes and Definitions

Our primary outcome was time to reach full enteral feeds, measured from the day when feeding was stopped due to NEC concern to the day infant could tolerate ³ 120 mL/kg/day of enteral feeds for 2 consecutive days, within 30 days of NEC concern. Secondary outcomes included time to end antibiotics and time to end bowel rest, measured from the day when antibiotics or bowel rest was started due to NEC concern until antibiotics or bowel rest was discontinued, within 14 days of NEC concern. Data were censored in the event of death or hospital transfer during the corresponding NEC episode. Safety outcomes included the frequency of death, surgical NEC, and the composite outcome of death or surgical NEC, as well as length of stay. Impact of imaging on decision-making was measured by a 5-point Likert-scale survey completed by treating neonatologists (**eTable 3 in the Supplement**).

### Sample Size and Data Analysis

Based on pilot data^25^ showing NEC ruled-out infants reached full enteral feeds in 5 ± 2.7 days, we estimated 29 infants per group (58 total) were needed to detect a 2-day difference with 80% power and a 2-sided α of 0.05. Assuming ∼40% of suspected NEC cases are ruled out and allowing for a 10% buffer for potential attrition, we projected enrolling 160 infants to meet the target sample size. Analyses followed the intention-to-treat principle, with a per-protocol sensitivity analysis excluding crossover. Primary and secondary outcomes were analyzed according to final diagnostic classification (*NEC Ruled In, Quick Rule-Out*, and *Extended Rule-Out*) as time-to-event variables using the Fine-Gray test for censoring and Cox proportional hazards (CPH) models for group comparisons. Infants who died or transferred were censored at the time of event. Subgroup analysis by NICU level (III vs IV) was conducted to assess for site-related differences. Safety outcomes were compared using chi-square or t-tests. Likert-scale survey responses were summarized as medians (IQR) and compared using pairwise Wilcoxon rank-sum tests. Statistical analyses were performed using R v4 (R Core Team, 2023) and SPSS® v29 (IBM Corp, 2021). Two-sided *P* < 0.05 was considered significant.

## RESULTS

### Participants

From September 2022 to December 2024, a total of 169 infants underwent 199 imaging evaluations for NEC concern (**Figure 1**). Demographic and clinical characteristics are shown in **Table 1**. About 84% were preterm, and 62% were recruited from the BUS-experienced level IV NICU. Baseline demographics did not differ between infants randomized to AXR only vs AXR + BUS. Of the 199 evaluations for NEC concern, 114 were assigned to AXR only and 85 to AXR + BUS (**Figure 1**). Thirty-one AXR-only evaluations also included BUS, and 3 AXR + BUS evaluations included only AXR, per attending discretion. Of the 199 NEC concern episodes, 66 were classified as *NEC Ruled In*, 63 as *Quick Rule-Out*, and 70 as *Extended Rule-Out*. Representative images of infant with *NEC Ruled In* and *Extended Rule-Out* are presented in **eFigure 1 and eFigure2 in the Supplement**, respectively.

**Figure 1.**
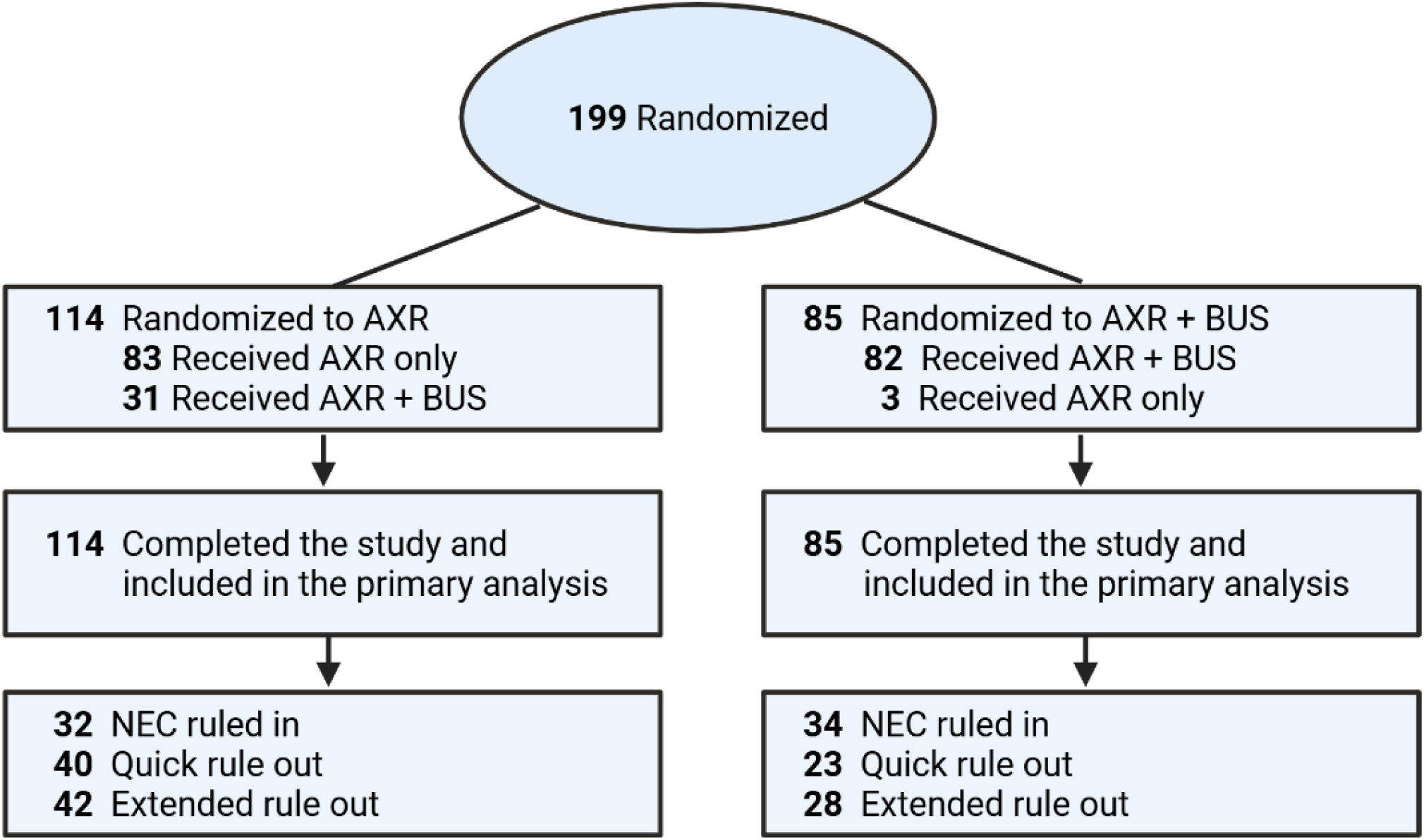
Participant Flow Diagram. A total of 199 infants were randomized to AXR (n = 114) or AXR + BUS (n = 85). Crossover occurred in 34 infants, with all participants completing follow-up and included in the primary intention-to-treat analysis. Infants were categorized as *NEC Ruled In*, *Quick Rule-Out*, or *Extended Rule-Out* based on clinical course.

**Table 1:**
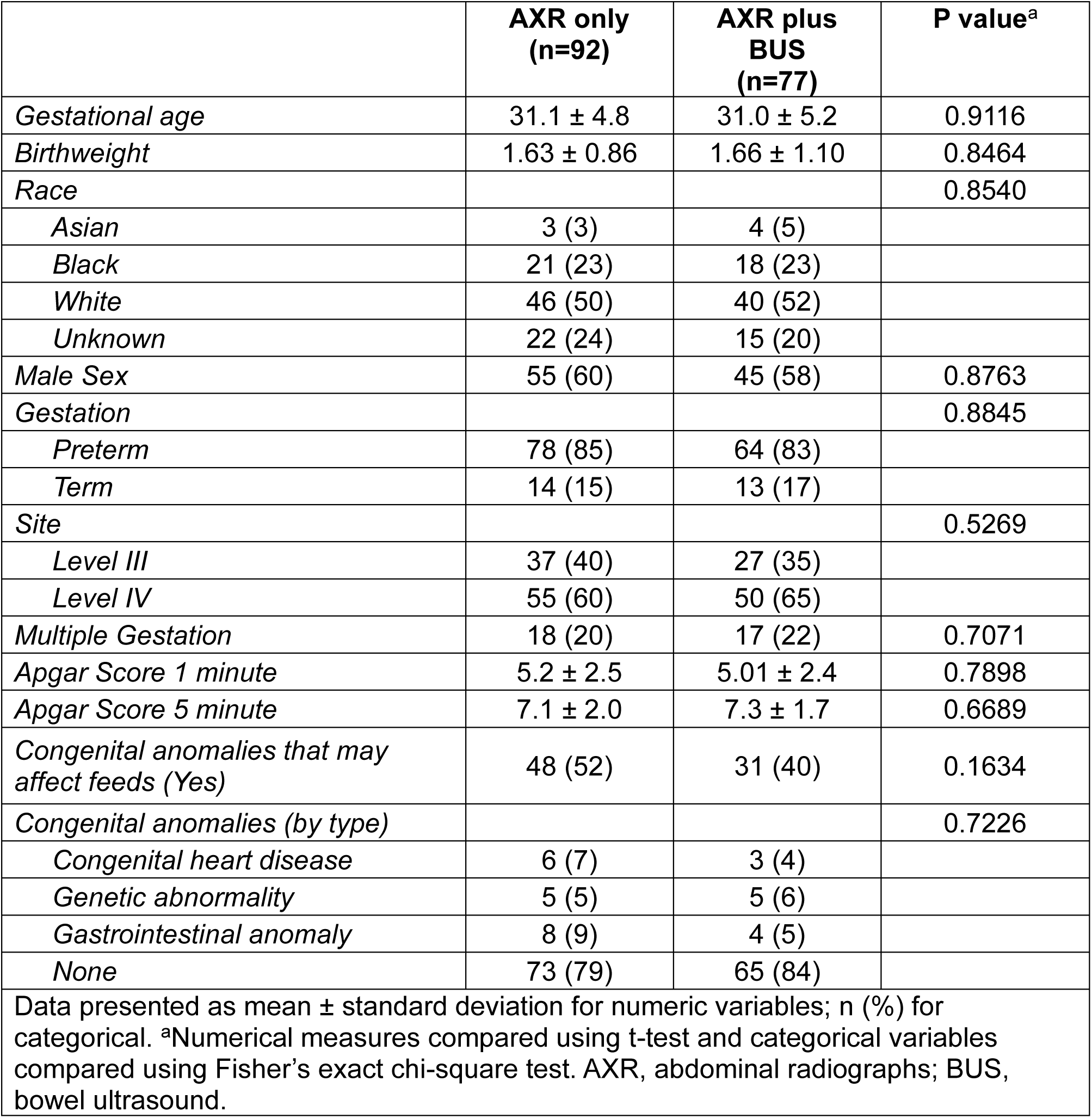
Baseline demographics of participants.

### Primary and Secondary Outcomes

Days to full feeds (*primary outcome*) did not differ between the AXR + BUS and AXR only groups for *NEC Ruled In* or *Quick Rule-Out* cohorts (**Table 2**). In contrast, infants with *Extended Rule-Out* reached full feeds sooner with AXR + BUS than AXR only (median 4 [IQR 1 – 5] days vs 8 [IQR 2 – 12] days, *P* = 0.009; **Table 2**). CPH analysis showed a higher likelihood of faster full feeds (hazards ratio [HR] 1.89; 95% CI 1.23 – 2.91) with adding BUS compared with AXR alone (**Table 2 and Figure 2**). Days to end bowel rest and antibiotics (*secondary outcomes*) were similar across imaging arms for all NEC classifications (**Table 2 and Figure 2**).

**Figure 2.**
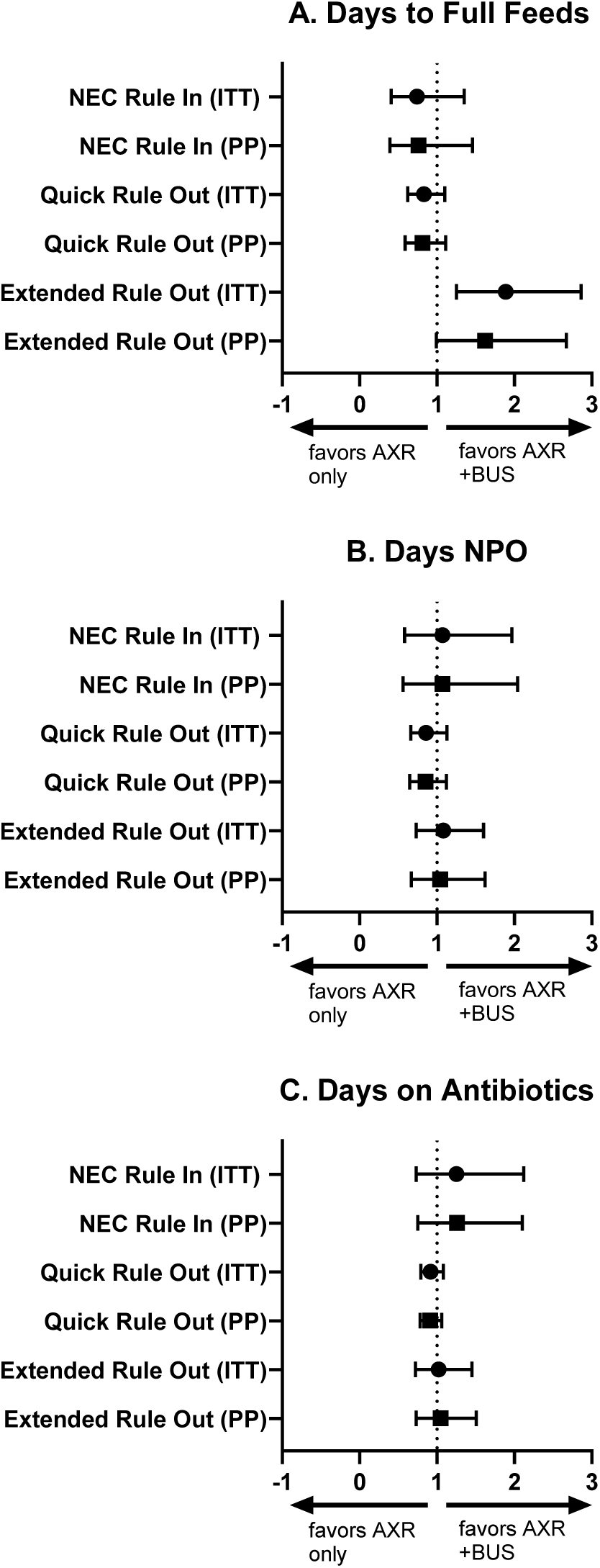
Forest Plot of Intention-to-Treat and Per-Protocol Analyses. Forest plot showing hazard ratios (HRs) and 95% CIs for the effect of adding BUS to AXR on time to full feeds (*primary outcome*), duration of bowel rest, and duration of antibiotics across NEC diagnostic cohorts. Separate estimates are shown for the intention-to-treat (circles) and per-protocol (squares) analyses. HRs greater than 1 favor BUS (faster time to event).

**Table 2:** Primary and secondary outcomes with intention-to-treat and with per protocol analysis.

|  | Intention to Treat Analysis |  |  |  |  |  | Per Protocol Analysis |  |  |  |  |  |
| --- | --- | --- | --- | --- | --- | --- | --- | --- | --- | --- | --- | --- |
|  | Median Time to Event (95% CI) |  |  | Cox Proportional Hazard Model |  |  | Median Time to Event (95% CI) |  |  | Cox Proportional Hazard Model |  |  |
|  | AXR only | AXR + BUS | P<sup>a</sup> | HR | 95% CI | P<sup>b</sup> | AXR only | AXR + BUS | P* | HR | 95% CI | P<sup>b</sup> |
| **NEC Ruled In** | **N=32** | **N=34** |  |  |  |  | **N=17** | **N=33** |  |  |  |  |
| *Days to full feeds* | 17 (11, 20) | 17 (14, 25) | 0.28 | 0.7<br>4 | 0.43 - 1.27 | 0.28 | 14 (10, 23) | 17 (12, 25) | 0.41 | 0.76 | 0.39 - 1.46 | 0.40 |
| *Days NPO* | 9 (7, 10) | 8 (7, 11) | 0.97 | 1.0<br>1 | 0.59 - 1.72 | 0.97 | 9 (5, 12) | 8 (7, 13) | 0.85 | 1.07 | 0.56 - 2.04 | 0.85 |
| *Days on Antibiotics* | 7 (7, 10) | 7 (6, 8) | 0.34 | 1.2<br>5 | 0.79 - 1.97 | 0.34 | 7 (7, 10) | 7 (6, 8) | 0.43 | 1.26 | 0.75 - 2.10 | 0.38 |
| **Quick Rule Out** | **N=40** | **N=23** |  |  |  |  | **N=38** | **N=21** |  |  |  |  |
| *Days to full feeds* | 0 (NA) | 0 (NA) | 0.29 | 0.8<br>3 | 0.62 - 1.11 | 0.20 | 0 (NA) | 0 (NA) | 0.28 | 0.81 | 0.59 - 1.11 | 0.19 |
| *Days NPO* | 0 (NA) | 0 (NA) | 0.40 | 0.8<br>7 | 0.67 - 1.12 | 0.28 | 0 (NA) | 0 (NA) | 0.39 | 0.85 | 0.65 - 1.12 | 0.25 |
| *Days on Antibiotics* | 0 (NA) | 0 (NA) | 0.18 | 0.9<br>2 | 0.80 - 1.05 | 0.22 | 0 (NA) | 0 (NA) | 0.17 | 0.91 | 0.78 - 1.06 | 0.22 |
| **Extended Rule Out** | **N=42** | **N=28** |  |  |  |  | **N=28** | **N=28** |  |  |  |  |
| *Days to full feeds* | 8 (2, 12) | 4(1, 5) | 0.00<br>9 | 1.8<br>9 | 1.23 - 2.91 | 0.00<br>3 | 6 (0, 10) | 4 (1, 5) | 0.08 | 1.62 | 0.99 - 2.67 | 0.05<br>7 |
| *Days NPO* | 2 (1, 3) | 2 (1, 2) | 0.31 | 1.2<br>4 | 0.83 - 1.84 | 0.29 | 1 (0, 3) | 2 (1, 2) | 0.88 | 1.04 | 0.67 - 1.62 | 0.87 |
| *Days on Antibiotics* | 0.5 (0, 2) | 0 (0, 2) | 0.68 | 1.0<br>7 | 0.76 - 1.50 | 0.70 | 0 (0, 2) | 0 (0, 2) | 0.81 | 1.05 | 0.73 - 1.51 | 0.80 |
| Data are shown as median time to event (95% confidence interval) estimated using the profile likelihood method. NA indicates non-estimable confidence interval due to all or most infants reaching the event on day 0. <sup>a</sup>P reflects unadjusted between-group comparison with Fine-Gray test; <sup>b</sup>P reflects adjusted comparisons with single predictor variable as intention to treat arm with the Cox model. AXR, abdominal radiographs; BUS, bowel ultrasound; HR, hazards ratio. |  |  |  |  |  |  |  |  |  |  |  |  |

To address crossover, we performed a per-protocol sensitivity analysis including only infants who received their assigned imaging intervention. Baseline demographics did not differ between those who did and did not receive their assigned imaging intervention (**eTable 4 in the Supplement**). Overall, results aligned with the intention-to-treat analysis (**Table 2** and **Figure 2**). For *NEC Ruled In* or *Quick Rule-Out*, no significant differences between imaging arms were observed. For *Extended Rule-Out*, BUS showed a trend toward shorter time to full feeds (HR = 1.62; 95% CI 0.99–2.67; *P* = 0.057), with direction and magnitude of effect similar to intention-to-treat analysis.

### Subgroup Analysis of Primary Outcome by Site

We assessed whether the effect of adding BUS on time to full feeds differed by site (**Table 3**), focusing on the *Extended Rule-Out* group because of small sample sizes in the other cohorts. In the intention-to-treat analysis, BUS shortened time to full feeds at the level IV NICU (median 4.5 vs 12 days; Fine-Gray *P* = 0.005) and showed a nonsignificant trend at the level III NICU (median 1 vs 2.5 days; Fine-Gray *P* = 0.275). CPH modeling confirmed that BUS accelerated time to full feeds overall (HR = 2.05, 95% CI 1.35–3.12) with no site interaction (*P* = 0.72). Per-protocol findings were directionally consistent: BUS remained associated with shorter time to full feeds at the level IV NICU (median 4.5 vs 9 days; HR = 1.79, 95% CI 1.09–2.93; *P* = 0.10), with no significant interaction by site (*P* = 0.55).

**Table 3.** Subgroup analysis by site of time to full feeds in extended rule out cohort.

| Subgroup | AXR only | AXR + BUS | Fine-Gray p<sup>a</sup> | Cox Model HR<sup>b</sup> | 95% CI | P for interaction<sup>c</sup> |
| --- | --- | --- | --- | --- | --- | --- |
| **Intention-to-Treat** | N=42 | N=28 |  |  |  |  |
| *Level IV* | 12 (2, 19) | 4.5 (3, 9) | 0.005 | 2.05 | 1.35 – 3.12 | 0.72 |
| *Level III* | 2.5 (0, 8) | 1 (0, 4) | 0.275 | -- | -- |  |
| **Per-Protocol** | N=28 | N=28 |  |  |  |  |
| *Level IV* | 9 (0, 19) | 4.5 (3, 9) | 0.10 | 1.79 | 1.09 – 2.93 | 0.55 |
| *Level III* | 2.5 (0, 9) | 1 (0, 4) | 0.19 | -- | -- |  |

Data presented as median days to reach full feeds (95% CI). <sup>a</sup>Fine-Gray p values indicate unadjusted comparisons between imaging arms within each subgroup. <sup>b</sup>Hazard ratio from the Cox proportional hazards model. <sup>c</sup>P for interaction = site x arm interaction from the Cox model. AXR, abdominal radiographs; BUS, bowel ultrasound; HR, hazards ratio.

### Safety Outcomes

Exploratory safety analyses for the first NEC concern episode are summarized in **eTable 5 in the Supplement**. In the intention-to-treat analysis, no significant differences were observed in the rates of death, surgical NEC, the combined outcome of death or surgical NEC, and length of stay. In the per-protocol analysis, surgical NEC was higher in AXR + BUS compared to AXR only (11% vs 1%; *P* = 0.03).

### Diagnostic and Therapeutic Thinking

Neonatologist likelihood ratings for diagnostic and therapeutic decisions after AXR and BUS are shown in **eTable 6 in the Supplement**. Ratings were based on a 5-point Likert scale, with higher scores indicating greater likelihood of performing the diagnostic or therapeutic action. For infants with *NEC Ruled In*, BUS increased the likelihood of diagnosing NEC (median 5 [IQR 4–5] vs 4 [2–4]; *P* = 0.001), initiating bowel rest (5 [5–5] vs 4 [4–5]; *P* = 0.019), and starting antibiotics (5 [5–5] vs 4 [2–5]; *P* = 0.003). In contrast, for Q*uick Rule-Out*, BUS was associated with a lower likelihood of bowel rest (1 [1–2] vs 2 [1–4]; *P* = 0.003) and ordering repeat imaging (1 [1–2] vs 2 [1–4]; *P* = 0.007). Similarly, for *Extended Rule-Out*, BUS decreased the likelihood of diagnosing NEC (2 [1–4] vs 3 [2–4]; *P* = 0.016), treating with bowel rest (4 [2-5] vs 4 [3-4]; *P* = 0.04), and ordering repeat imaging (4 [2–5] vs 5 [4–5]; *P* = 0.004).

## DISCUSSION

In this randomized diagnostic trial, we assessed the impact of adding BUS to standard AXR for infants undergoing evaluation for suspected NEC. Among infants with *extended rule-out* evaluations – those requiring repeated imaging but ultimately without NEC – adding BUS accelerated return to full feeds. Adding BUS also did not prolong bowel rest or antibiotic use and did not increase adverse outcomes, including surgical NEC, death, or length of stay. Moreover, neonatologists reported greater diagnostic and therapeutic confidence when BUS was incorporated.

Most prior studies of BUS for suspected NEC have been small, single-center cohort studies that retrospectively evaluated its use after adoption into the diagnostic pathway.^19–23^ Such studies are confined to diagnostic accuracy (Level 2) in Fryback and Thornbury’s hierarchy of diagnostic efficacy. Building on our pilot study,^25^ which demonstrated the feasibility of a randomized diagnostic trial, the current study represents the first prospective, multi-site randomized evaluation of BUS for NEC that addresses higher levels of diagnostic efficacy. Our prospective investigation allowed us to survey neonatologists regarding the impact of adding BUS on their diagnostic thinking (Level 3) and therapeutic decision-making (Level 4). Moreover, the randomized study design allowed us to determine the effect of adding BUS on patient outcomes (Level 5). Our study thus elevates the evidence for BUS beyond diagnostic accuracy (Level 2), demonstrating its value in shaping clinical decisions and improving patient outcomes (Levels 3–5).

Because most prior BUS studies for NEC were limited to level IV NICUs in freestanding children’s hospitals, questions remain about its generalizability to other settings.^32,33^ To address this, our trial included a level III NICU within an adult hospital, where imaging was performed by adult sonographers and radiologists with no prior experience using BUS for NEC. Subgroup analysis by site showed a consistent direction of effect: adding BUS was associated with shorter time to reach full feeds in both level III and level IV NICUs. The magnitude of reduction was also similar – about half the time to full feeds in each subgroup – but only the level IV site reached statistical significance, likely due to smaller sample size at the level III site. Of note, infants at the level IV site had longer overall time to reach full feeds than those at the level III site, possibly reflecting differences in feeding practices or greater illness complexity in higher-acuity level IV NICU.^34,35^ Nevertheless, despite their differences, the similar direction and magnitude of effect observed at both sites provide supporting evidence that adding BUS to AXR can improve clinical outcomes even in adult hospital settings without prior BUS expertise.

The improvement in feeding recovery among infants with *Extended Rule-Out* evaluations likely reflects how BUS informs clinical decision-making. BUS provides real-time visualization of bowel perfusion, peristalsis, and wall integrity – added information that may reduce diagnostic uncertainty.^36^ For example, prior studies show how BUS can detect early disease features not yet apparent on AXR and offer reassurance when findings are normal.^18,37,38^ In contrast, among infants with *Quick Rule-Out* evaluations – those in whom NEC was excluded after a single AXR – adding BUS did not seem to confer additional benefit. This finding suggests that BUS is most valuable when diagnostic uncertainty persists and repeated imaging is required, rather than in clear-cut cases easily ruled out by initial AXR. Among infants with *NEC Ruled In*, we observed no significant differences between imaging arms in any of the primary or secondary outcomes. This finding likely reflects the standardized approach to management once NEC is diagnosed, as bowel rest and antibiotic duration are typically uniform based on modified Bell staging.^30,31^

The higher rate of surgical NEC in the BUS arm – statistically significant in per-protocol but not in intention-to-treat analysis – warrants careful interpretation. We hypothesize that BUS enables earlier and more accurate recognition of advanced disease that facilitates surgical intervention.^16,20,22^ Unlike AXR, which primarily relies on free air to indicate need for surgery, BUS offers additional markers of advanced disease – such as complex ascites, loculated fluid collections, bowel wall thinning, and absent or diminished bowel perfusion – which may influence surgical decision-making.^11,12,16^ The difference between intention-to-treat and per-protocol analyses likely reflects the impact of crossover. Although the exact rationale was not documented, we surmise that infants who crossed over – most of which were from AXR to BUS – were perceived by clinicians as clinically sicker, prompting the decision to obtain additional information from BUS. In the intention-to-treat analysis, these infants remained in the AXR group to preserve randomization, which may have reduced the observed effect of BUS.^39^ In contrast, the per-protocol analysis excluded these infants, which may potentially introduce selection bias by removing sicker infants and amplifying the observed difference between groups.^40,41^ Overall, these findings highlight the potential importance of BUS in surgical decision-making and underscore the need for future studies to determine whether BUS genuinely improves surgical outcomes.

Although not powered, the use of BUS did not appear to compromise safety, as mortality rates were similar between study arms. Moreover, recurrent NEC occurred less frequently in the AXR + BUS arm, possibly due to greater diagnostic clarity with BUS compared to AXR alone.^38^ Surgical NEC did occur more frequently in the BUS arm, a finding that may reflect earlier or more accurate recognition of advanced disease with BUS, as discussed above. The modest trend toward longer hospital stays among infants randomized to BUS also likely reflects the higher proportion of surgical NEC in this group. Larger studies are needed to determine whether BUS truly improves timeliness and appropriateness of surgical intervention for NEC while maintaining safety and reducing recurrent disease.

### Limitations

We acknowledge several limitations. First, the study’s sample size limited statistical power, particularly for precise subgroup analyses across NICU levels and diagnostic cohorts. Second, the unblinded design and occasional crossover could have introduced bias to the study. Third, although imaging protocols were standardized, variations in sonographer expertise and local practices could have contributed to site-level differences. Fourth, our study included only one level IV and one level III NICU, which limits the generalizability of our findings. Fifth, primary and secondary outcomes focused on short-term clinical course and surrogate markers of disease and recovery; larger multicenter studies should evaluate more meaningful outcomes such as death, short gut, and neurodevelopment. Finally, this trial did not examine cost-effectiveness of BUS, corresponding to Level 6 (societal benefit) in Fryback and Thornbury’s hierarchy of diagnostic efficacy.^24^

## Conclusion

This randomized diagnostic trial provides the first prospective evidence that adding BUS to standard AXR can influence clinical decision-making and improve outcomes for infants evaluated for NEC. BUS appears particularly valuable in infants with prolonged diagnostic uncertainty, where it can reduce unnecessary treatment and support earlier feeding recovery, without increasing mortality or overall adverse events. Larger multicenter comparative-effectiveness studies are needed to confirm these findings, determine which patient populations benefit most from BUS, and identify strategies for its effective and sustainable implementation across diverse NICU settings.

## Data Availability

De-identified data that support the findings of this study are available from the corresponding author upon reasonable request.

## Conflict of Interest Disclosures

All authors have completed and submitted the ICMJE Form for Disclosure of Potential Conflicts of Interest. No disclosures were reported.

## Funding

This study was supported by the American College of Radiology Fund for Collaborative Research in Imaging Grant

## Notes

### Competing Interest Statement

The authors have declared no competing interest.

### Clinical Trial

ClinicalTrials.gov Identifier NCT05573113

### Author Declarations

The IRB of Children's Mercy Hospital gave ethical approval for this work.

